# How long do nursing staff take to measure and record patients’ vital signs observations in hospital? A time-and-motion study

**DOI:** 10.1101/2020.12.09.20246355

**Authors:** Chiara Dall’Ora, Peter Griffiths, Joanna Hope, Jim Briggs, Jeremy Jones, Stephen Gerry, Oliver Redfern

## Abstract

**Introduction:** Monitoring vital signs in hospital is an important part of safe patient care. However, there are no robust estimates of the workload it generates for nursing staff. This makes it difficult to plan adequate staffing to ensure current monitoring protocols can be delivered.

**Objective:** To estimate the time taken to measure and record one set of patient’s vital signs observations; and to identify factors associated with time to measure and record one set of patient’s vital signs observations.

**Methods:** We undertook a time-and-motion study of 16 acute medical or surgical wards across four hospitals in England. One hospital recorded vital signs on paper, while three recorded measurements on electronic devices. Two trained observers followed a standard operating procedure to record the time taken to measure and record vital sign observations. We used mixed-effects models to estimate the mean time using whole observation rounds, which included preparation time, or time spent taking observations at the bedside. We tested whether our estimates were influenced by nurse, ward and hospital factors.

**Results:** After excluding non-vital signs related interruptions, dividing the length of an observation round by the number of observations in that round yielded an estimated time per observation set of 5 minutes and 1 second (95% Confidence Interval (CI) = 4:39-5:24). If interruptions within the round were included, the estimated time was 6:26 (95% CI = 6:01-6:50). If only time taking each patient’s observations at the bedside was considered, after excluding non-vital signs related interruptions the estimated time was 3:45 (95% CI = 3:32-3:58). We found no substantial differences by hospital, ward or nurse characteristics, despite different systems for recording observations being used across the hospitals.

**Discussion:** The time taken to observe and record a patient’s vital signs is considerable, so changes to recommended observation frequency could have major workload implications. Variation in estimates derived from previous studies may, in part, arise from a lack of clarity about what was included in the reported times. We found no evidence that nurses save time when using electronic vital signs recording, or that the grade of staff taking the observation influenced the time taken.

**Conclusions:** Taking and recording vital signs observations is time consuming and the impact of interruptions and preparation away from the bedside is considerable. When considering the nursing workload around vital signs observations, no assumption of relative efficiency should be made if different technologies or staff groups are deployed.

## BACKGROUND

Monitoring vital signs in acute hospitals (e.g. blood pressure, heart rate, and temperature) is an important part of ensuring safe patient care (Odell et al., 2009). Acutely unwell patients have their vital signs measured regularly (Brekke et al., 2019), as abnormalities in vital signs can indicate that a patient’s condition has deteriorated (Churpek et al., 2016, Kause et al., 2004). Failure to identify deterioration early is associated with worse patient outcomes and contributes to avoidable deaths (National Institute for Health Care Excellence, 2007, National Confidential Enquiry into Patient Outcome and Death and 2015, Keogh, 2013, Hogan et al., 2012). Nurses often report missing essential patient surveillance due to high workload: one recent study found that around 35% of the vital sign observations scheduled according to an early warning score based protocol were delayed or missed (Redfern et al., 2019, Ball et al., 2014). However, the uncertainty around appropriate monitoring frequencies, and the impact these have on nursing workload, makes accurate workforce planning challenging (Smith et al., 2017, Dall’Ora et al., 2020a).

Outside of high acuity areas, such as Intensive Care and High Dependency Units, vital signs are rarely monitored continuously. Instead, clinical staff (usually nurses), measure and record patients’ observations at regular intervals, often guided by protocols (Rose and Clarke, 2010). Protocols vary widely across different healthcare systems and are usually based on expert consensus (Smith et al., 2017). For example, in the US, many medical units default to observing all patients every four hours (Orlov and Arora, 2020, Tham et al., 2020). In many Northern European countries, monitoring is usually guided by protocols based on Early Warning Scores (EWS), which combine individual vital sign abnormalities into a single score. For example in the UK, the National Early Warning Score 2 (NEWS2) system is recommended by national bodies for use in acute hospitals and has been implemented in most settings (Royal College of Physicians, 2017, National Institute for Health Care Excellence, 2007, National Confidential Enquiry into Patient Outcome and Death and 2015). Patients with a low NEWS2 are monitored every 6-12 hours, increasing to hourly for patients with scores above 6 (Royal College of Physicians, 2017).

Routine four-hourly observations can result in unnecessary observations and have been recommended for de-implementation (Orlov and Arora, 2020, Tham et al., 2020). However, determining alternative observation regimens remains problematic. Although there is evidence that EWSs are good predictors of adverse patient outcomes and so could be used to prioritise patients most in need of frequent observation, there is no direct evidence to link these scores to specific monitoring frequencies (Smith et al., 2017). Protocols based on EWSs can generate high numbers of false alerts, unnecessarily subjecting stable patients to high frequency monitoring, and potentially generating unnecessary work for nurses (Bedoya et al., 2019, Fox and Elliott, 2015). There is growing evidence that adherence to EWS-based monitoring protocols is poor (Hands et al., 2013, Tysinger, 2014, Redfern et al., 2019). The perception that many observations are unnecessary in low-risk patients may be one of the underlying causes of poor adherence (Redfern et al., 2019), with staff reporting they have to prioritise some tasks such as responding to rapid deterioration over scheduled observations (Hope et al., 2019). New technology aiming to improve the fidelity of vital signs monitoring, including wearable devices and even providing patients with devices to measure and record their own vital signs (Weenk et al., 2018), promises considerable time savings for nursing staff (Wong et al., 2017, Bellomo et al., 2012, Wager et al., 2010).

However, while these different approaches promise a more efficient use of nursing time with potential benefit to patients, there is currently little data to show how much time nurses spend on vital signs activities or to estimate potential savings (or costs) associated with change. A recent literature review found limited evidence with considerable variation between and within studies, 3 concluding that such uncertainty means that existing evidence cannot reliably inform workload planning for vital signs observations (Dall’Ora et al., 2020a). Limitations of existing research included lack of standardisation in measuring vital signs, and inadequate description of the observation methods used. Factors such as the influence of interruptions to work, the time required to prepare and source equipment and the expertise of the staff involved were not considered.

The aim of this observational study was to estimate how much time nursing staff expend measuring and recording a set of vital signs observations in general hospital wards across four English hospitals; and to identify factors influencing the time involved with measuring and recording vital signs observations.

## METHODS

### Design and setting

This was a time-and-motion observational study, conducted in 16 inpatient adult general wards within four acute NHS hospitals in the south of England.

### Sample and recruitment

Hospitals were selected a priori within the South East region in order to get variation in the approaches to vital signs recording. All hospitals used NEWS2 to guide the frequency at which patients are observed, as this is now recommended for use in all acute hospitals in England. Three hospitals used electronic systems to record vital signs. These electronic systems differed in each hospital, but they shared the embedded algorithm to calculate NEWS2 scores. In one hospital, vital signs were recorded on paper charts at the patient’s bedside and the NEWS2 was calculated manually.

There was little existing data to help calculate the required sample size and estimation was further complicated by clustering of observations in nurses and units. As a guide, we used the confidence intervals reported by Wong and colleagues (Wong et al., 2017), a study with a similar clustering structure to our planned study, to provide an estimate. Based on their 95% confidence intervals, we inferred a standard error of 10.96 (standard error = 95% CI/(2*1.96)). Based on their standard error, in a sample of 280 observations from wards with electronic recording of vital signs, we calculated that a sample of 640 sets of observations would give an estimated mean with a precision of +/-<10% if each set of observations took three minutes. For each ward, we planned to observe for a total of eight hours, spread across four sessions, aiming to observe a minimum of 10 sets of vital signs measurements per session.

In each study hospital, we identified general medical/surgical inpatient wards that met our inclusion/exclusion criteria. Inclusion criteria were: ward admitting adult patients (i.e. 18 years old or above) only. Wards occasionally admitting adolescents of younger were are eligible; wards open seven days a week with most patients having overnight stays of at least one day; wards with patients requiring Level 0 and Level 1 care^1^ We then randomly selected four wards. If a ward manager did not agree to participate, we randomly selected another ward. In each ward, the ward manager provided consent for the research on behalf of the ward staff, and we sought to recruit all nursing staff (i.e. registered nurses, healthcare assistants, nursing associates, student nurses) working on the ward at the time of each observation session. Staff were given the chance to opt out of the research when the observation session began and when approached during the session. Patients and relatives were informed about the study through posters displayed on the ward and through explanation given by the researchers and/or nursing staff when patients were having their vital signs monitored. We explained to patients that they were not the focus of our studies and that we were observing nursing staff only, and gave them the option to opt out in case they did not want to have researchers near their bed space, which only occurred twice.

### Data collection

Data were collected using the Quality of Interactions (QI) Tool, a bespoke software on an Android tablet that enables users to enter data in real-time. The QI tool also enabled us to collect contextual data, including the total number of patients on the ward, and numbers of registered nurses (RNs), nursing assistants and student nurses on shift during the observation session. More details on the tool can be found at (Bridges et al., 2018). Two trained researchers conducted observations as non-participant observers. At the end of each session, researchers asked staff whether they had modified their practice as a result of being observed. The definition of vital signs activities recorded during observations are reported in Table 1.

**Table 1.** Definitions of activities related to vital signs.

|  |  |
| --- | --- |
| Vital sign | One or more of the six physiological parameters that form the basis of the NEWS2 scoring system (Royal College of Physicians, 2017): respiration rate, oxygen saturation, systolic blood pressure, pulse rate, temperature and level of consciousness or new confusion. |
| Vital signs equipment | Sphygmomanometer, manual blood pressure cuff, stethoscope, oxygen pulse oximeter, thermometer, electronic recording system for vital signs, tablets. |
| Vital signs documentation | Observing previous vital signs trends electronically or on paper, patient notes, track and trigger charts, other vital signs observation charts. |
| Vital signs observation set | The collected acts of taking and recording the measurements of vital signs and sourcing vital signs equipment per patient. These can be complete (i.e. there is no reason for the observer to suppose that the six physiological parameters of vital signs as outlined in NEWS2 have not all been completed) or incomplete (i.e. it is obvious to the observer that not all six physiological vital signs have been completed). |
| Vital signs round | One or more sets of individual vital signs observations taken sequentially by a single nurse are clustered in a vital signs round. Captures all parts of taking vital signs observations including using 'Vital signs documentation' and 'Vital signs equipment' as well as carrying out 'Vital signs observation sets'. |
| Interruption | An act outside the process of 'Vital signs', and 'Vital signs documentation' – see "Vital signs related interruption" and "Non-vital-signs related interruption" for more information. |
| Vital signs related interruption | Interruptions that would not have occurred unless vital signs were being taken (e.g. cleaning vital signs monitoring equipment, sourcing alternative equipment when equipment was faulty, |
|  | escalation to other staff members if vital signs were abnormal). |
| Non-vital-signs related interruption | Interruptions that are outside of the process of 'Vital signs round', for example the provision of personal care; the nurse leaving the patient to speak to someone else or to attend to other patients. |

We established criteria for start and stop times of the above activities. We were interested in the preparation time occurring at the beginning of the observations round, as this is work associated with vital signs, and we wanted to take interruptions into account if they contained any vital-signs related activity. In summary, a vital signs round was deemed to start every time nursing staff sourced vital signs equipment or vital signs documentation, and to finish when one or more sets of vital signs observations were taken and the vital signs equipment and/or documentation were replaced. A vital signs observation set was deemed to start when a nurse entered the bed space and measured one or more of the six physiological parameters of the NEWS2 scoring system, and to end when the nurse left the bed space and the measuring of vital signs as defined by NEWS2 had finished (see (Dall’Ora et al., 2020b) for more detail). For our definition of vital-signs vs non vital-signs related interruptions, see Table 1.

Observers were trained using an observation guide. The guide was found to be easy to use and led to a high level of inter-rater agreement, with a mean difference between raters of only 3 seconds per set of vital signs observation (mean observation estimate 3 minutes 47 seconds) and limits of agreement from + 19 to −13 seconds (Dall’Ora et al., 2020b). The cumulative difference over 4 hours of observation during the training sessions was less than 2% of the total time observed.

### Data management

After each observation session, data were uploaded onto the QI Tool server. This server and the associated databases were located at and managed by the IT team of the raters’ institution. We did not store any personal data. Before data analysis, one member of the team checked all data extracts for any errors: for example, if an observer had indicated that the timing of an interaction was incorrect (for example where there had been an inadvertent delay in recording the end of the observation). These instances were rare (n=37) and were corrected, and all interruptions were coded as vital signs related or not vital signs related.

### Statistical analysis

Our data comprised sets of vital sign observations within observation rounds. As well as the time taken to measure and record vital signs, each round included preparation time and interruptions. For this reason, we estimated the time taken to perform a set of vital sign observations in three ways:

1. Our first estimate (E1) was calculated by dividing the length of the round by the number of vital signs observation sets.
2. Our second estimate (E2) was the time taken at the patient’s bedside, between when the nurse entered and left the bed space.
3. Variants of the E1 and E2 estimates were calculated by removing time associated with some or all interruptions (e.g. non vital signs related such as discussions with relatives).

To account for heterogeneity within hospitals and wards, we used mixed-effects models with nested random effects terms for wards within each study hospital. To test whether the time taken to obtain observations was influenced by nursing staff grade (i.e. registered nurse, nursing assistant and student nurse) or study hospital, we fitted a second model where nursing staff grade and study hospital were set as fixed effects. Coefficients from these models were used to estimate adjusted (conditional) means for each hospital and staff grade. Model-based estimates were compared to raw means, with confidence intervals calculated using bootstrapping (2000 samples).

To determine whether any efficiency was gained by measuring individual observations within a round, we fit a mixed model to estimate the total time of the round, with the number of observations as a fixed effect and nested random effects terms for wards within each study hospital. Conditional means from this model allowed us to estimate the marginal effects from changing the length of rounds.

All analyses were performed with R version 4.0 (R Development Core Team, 2020) using the glmmTMB package for mixed-effects models (Brooks et al., 2017) and the emmeans package to calculate conditional means (Lenth et al., 2020).

### Ethical approvals

We obtained ethical approval for this study from the Research Ethics Committee, South Central – Berkshire, Committee ref: 19/SC/0190.

## RESULTS

We undertook 64 sessions in four hospitals. In hospitals 1, 2, and 3 vital signs observations were recorded on electronic devices. Hospital 4 recorded vital signs observations on paper charts after which nurses manually calculated the NEWS2. In total, we observed 715 sets of vital signs measurements (181 in Hospital 1, 219 in Hospital 2, 140 in Hospital 3, and 175 in Hospital 4). Of these, 680 (95%) were complete sets where all six vital signs were measured and recorded. These observations were clustered in 260 rounds, with a median of 2 observations per round (interquartile range: 1-4; range: 1-11).

Registered nurses performed 355 observations (50%) nested in 122 rounds (47%). Healthcare assistants performed 217 observations (30%) nested in 81 rounds (31%). Student nurses performed 143 observations (20%) nested in 57 rounds (22%). There was little evidence that observation changed behaviours, although in five sessions, a staff member reported they felt their behaviour had changed as a result of being observed. Table 2 displays the number and proportions of vital signs sets performed by staff group per hospital.

**Table 2.** Number and proportions of vital sign sets by staff group per hospital

| Number of vital sign sets | Hospital 1<br>N (%) | Hospital 2<br>N (%) | Hospital 3<br>N (%) | Hospital 4<br>N (%) | Total |
| --- | --- | --- | --- | --- | --- |
| Registered Nurses | 54 (30%) | 163 (74%) | 116 (83%) | 22 (13%) | 355 |
| Healthcare assistant | 63 (35%) | 32 (15%) | 1 (1%) | 121 (69%) | 217 |
| Student nurse | 64 (35%) | 24 (11%) | 23 (16%) | 32 (18%) | 143 |
| Total | 181 (100%) | 219 (100%) | 140 (100%) | 175 (100%) | 715 |

The proportion of vital signs taken by registered nurses ranged from 13% in Hospital 4 to 83% in Hospital 3. In hospitals 2 and 3, registered nurses took the majority of the vital signs observations, whereas healthcare assistants and student nurses did so in hospitals 1 and 4.

When considering estimates from rounds, where we calculated the time as total round length divided by the number of observations within that round, the mean time per observation set excluding non-vital-signs related interruptions was 5:01 (95% CI = 4:39-5:24).

When considering the time at the patient bedside only (i.e. vital signs observation sets without the preparatory time that is shared out across a round), the mean time after excluding non-vital-signs related interruptions was being 3:45 (95% CI 3:32-3:58). Table 3 summarises all marginal means from the mixed effects models. The raw estimates did not differ substantially from the mixed effects models’ estimates, and can be found in Supplementary File 1. There was some positive (right) skew in the distribution of time taken, with means slightly higher than the median although the distributions were not grossly asymmetrical (Supplementary File 1.figure s2).

**Table 3.** Marginal means time per observation set from the mixed effects models

| **Including Interruptions**<br>**(minutes:seconds),**<br>**mean [95% CI]** | **Excluding**<br>**Non Vital Sign Interruptions**<br>**(minutes:seconds), mean [95%**<br>**CI]** | **Excluding**<br>**All Interruptions**<br>**(minutes:seconds),**<br>**mean [95% CI]** |
| --- | --- | --- |
| **From rounds** |  |  |
| 6:26 [6:01-6:50] | 5:01 [4:39-5:24] | 4:15 [4:04-4:27] |
| **At the patient bedside only** |  |  |
| 4:24 [4:11-4:38] | 3:45 [3:32-3:58] | 3:31 [3:22-3:40] |

Differences between hospitals and staff groups were small and there was no substantial variation associated with wards. Table 4 shows the marginal mean estimates (excluding non-vital signs observations) for staff groups and hospital. Round based estimates ranged from 4:53 for registered nurses to 5:16 for student nurses. For hospitals, estimates ranged from 4:38 per set of observations to 5:41. In both cases there was substantial overlap between the confidence intervals. Differences based on bedside only observations were much smaller.

**Table 4.**
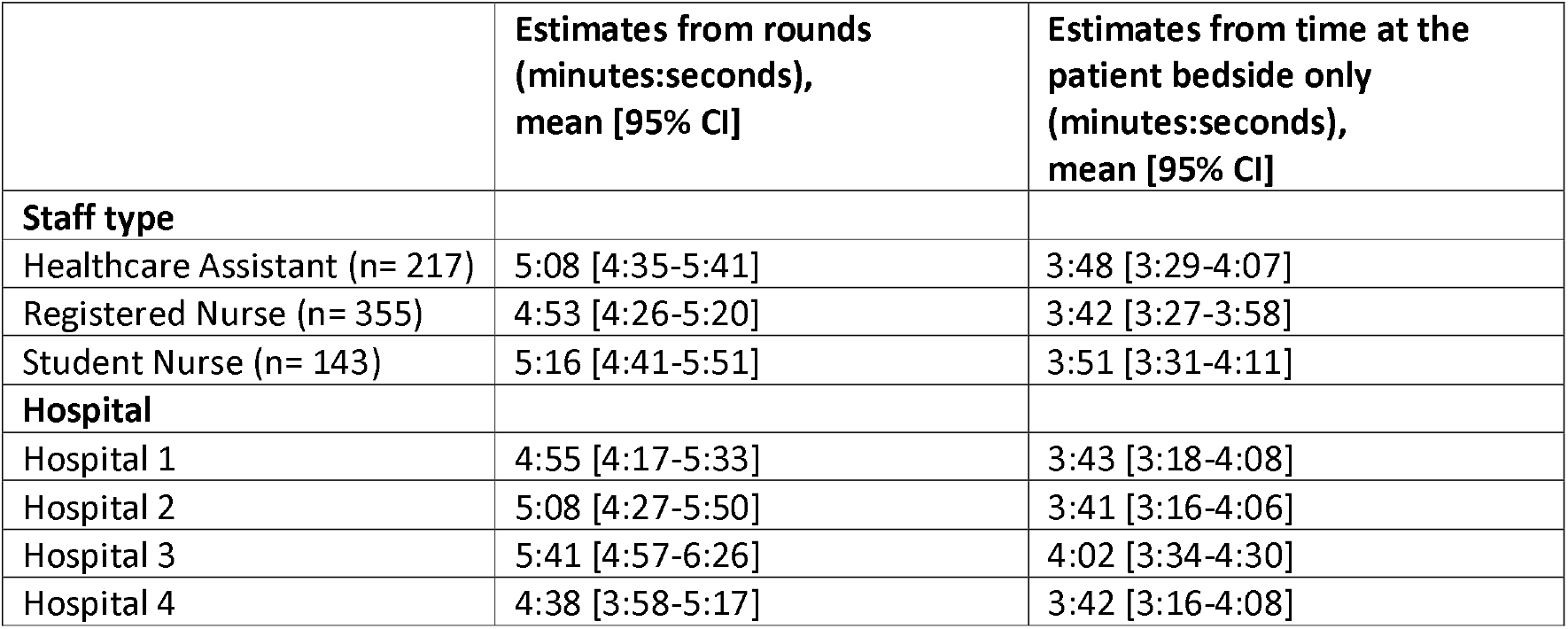
Marginal means from mixed effect models by staff group and hospital

When considering efficiencies deriving from grouping observations of multiple patients in the same round, we found substantial efficiency gains from undertaking observations as part of a round as opposed to observing a single patient (effectively a round comprising one set of vital signs observations). However, marginal gains from longer (as opposed to shorter) rounds were low. Using estimates excluding non-vital signs observations, we estimated that a round where two patients are observed takes 7% less time per person observed than observing a single patient. This occurs because preparatory time such as sourcing equipment and travel to the patient care area is divided across more than one patient. Marginal gains reduce as round length increases. A round where five patients are observed takes 12% less time per patient compared to observing a single patient while rounds of 10 patients give a time saving of 13% per patient (see Figure 1).

**Figure 1.**
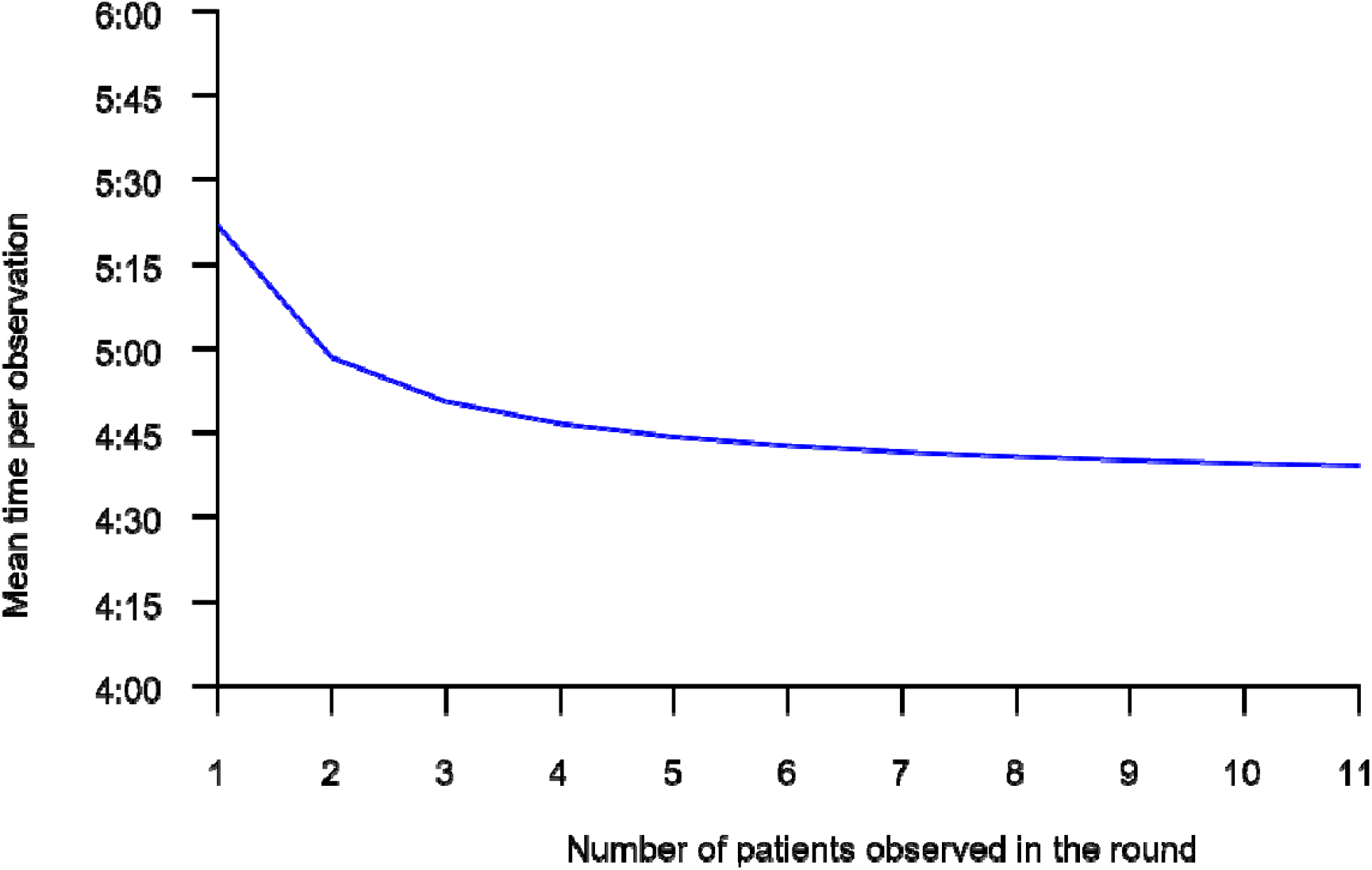
Time per set of observations Vs number of patients observed in a round

## DISCUSSION

After removing non-vital-signs related interruptions, we estimated that a single set of vital sign observations lasts, on average, three minutes and forty-five seconds when observing time spent at the bedside. By considering observations as part of a round as opposed to simply observing the work at the bedside, this estimate increased to 5:01. The estimated time varied substantially depending on how interruptions were treated.

Previous estimates of the work involved in taking vital signs observations has generally been on a smaller scale and has paid scant attention to issues such as interruptions or activities associated with preparation. Our estimates for taking an individual set of vital signs are broadly similar to those reported by Wong and colleagues (Wong et al., 2017), who estimated a geometric mean of 3:58 for taking and recording vital signs observations on paper. However, our results show that the preparation time, which is included in our round-based estimates, can be substantial. Our higher estimate of 5:01 per set of vital signs, based on averaging the round time while excluding non-vital sign interruptions, is likely to be a more realistic estimate of the work involved. Using this estimate, our findings are closer to those of Clarke (Clarke, 2006) and Zeitz (Zeitz, 2005) who both estimated 5 minutes and 40 seconds.

Our results show that efficiencies are potentially gained from clustering observations in rounds: although individually tailored observation schedules, as occurs when using an early warning score-based protocol, mitigate against this. Although a tailored approach may avoid unnecessary observations, there are likely to be efficiency gains from undertaking rounds because the ‘overhead’ of preparation is divided between more patients. However, the efficiency savings from longer rounds are small, with most efficiency gains achieved with rounds involving the observation of 2-4 patients and little additional benefit from rounds of more than 5.

Studies have reported substantially lower times when recording vital signs observations electronically, compared to pen and paper (Wong et al., 2017, Yeung et al., 2012, Bellomo et al., 2012). By contrast, we did not find a large difference between the three hospitals that used electronic devices for recording and the one that used paper charts. Indeed while between hospital differences were small, the hospital that used paper charts (hospital 4) provided the shortest mean estimate for time taken. Previous studies comparing paper and electronic observations have deployed a before and after design, where data were collected shortly after electronic systems were implemented. In our study, we collected data in settings where vital signs processes and systems had been well-established and routinised, so that the estimates we found might better reflect normal practice, and may indicate that any time savings achieved when the technology was first introduced might reduce with time. Early performance improvements such as this could be an example of a “Hawthorne effect” whereby research participants increase their productivity because they are being observed while using a new technology framed as having the potential to save time and reduce workload (Wickström and Bendix, 2000). Certainly, this is a useful reminder that no efficiency assumptions should be made when introducing new technologies to reduce nurses’ workload, and evidence offered of short-term benefits should be treated with caution because any initial time savings might dissipate as the technology becomes normalised and observation ceases.

We did not find the time taken to undertake vital signs observation varied between the staff groups taking the observations. Vital signs observations are a fundamental nursing activity (Rose and Clarke, 2010), but there is evidence that hospitals have different policies regarding which staff group is mainly responsible for vital signs observations. For example, in Hospital 4 of the study, the majority of vital signs observations were undertaken by healthcare assistants, while in Hospital 3 almost all were undertaken by registered nurses. The issue of delegation in nursing is a complex one (Potter et al., 2010). Although we found that all staff groups appear equally efficient at taking vital signs, concerns have been raised about the ability of assistant staff to properly interpret and act on vital signs and to fully assess the wider patient condition (Mok et al., 2015). Given the work involved in supervising and delegating and the potential to lead to adverse outcomes even though the task itself is completed efficiently (Griffiths et al., 2018), any apparent economic benefits associated with substitution of lower paid staff may be illusory.

Given the uncertainty associated with the currently recommended observation frequencies, including those associated with Early Warning Scores, there is considerable potential for change which may involve either increased or decreased monitoring. Such changes have significant implications for workload and the results of this study provide an important tool for quantification. Vital signs observation on a 30 bedded unit on which all patients are observed every 4 hours would require over 900 minutes of nursing time per day (based on our round based estimate excluding non vital signs observations). The workload associated with changes in vital signs observations must be considered alongside the clinical benefits of those changes in order to ensure there are adequate resources and/or that benefits from releasing nurses to deliver other care are realised. More recently, continuous vital signs monitoring has been proposed as a solution to avoid missing patient deterioration while also reducing nurses’ workload (Downey et al., 2018, Sun et al., 2020). Studies such as this provide some guide as to the potential time savings of implementing continuous vital signs systems, but it would be unwise to assume that such systems reduce the requirement to zero. Consideration needs to be given to the ancillary value that arises from the incidental checking and important conversations with patients that occur alongside taking the vital signs. While we observed considerable time taken up by ‘interruptions’, these interruptions may represent opportunities taken to perform other valuable work and removing the vital signs observation could remove the opportunity.

### Limitations

We observed only on week days between 9 AM to 5 PM. However, there is no a-priori reason to assume that time taken differs at night, although this is possible due to increased time taken to wake patients up and/or the challenges associated with measuring and recording vital signs observations in low light conditions. Our sample is also based on UK hospitals only so our results may not generalise to other countries, however our sample is bigger than most previous studies, which have been mainly conducted in a single hospital (Adomat and Hicks, 2003, Clarke, 2006, McGrath et al., 2019). In common with other studies the presence of observers and awareness of being observed might have influenced the behaviours of staff, although we explained carefully to participants that the aim of the study was to achieve a realistic picture of vital signs observations and not to measure their performance. In most cases staff reported that they did not feel observation influenced behaviour although we cannot discount unconscious changes. The direction of any resulting bias is unpredictable as it could lead to either slower (more careful / diligent) observation or faster (more focussed / efficient) performance. We have presented arithmetic means because our main purpose is to give an estimate of the work involved which can be summed across activities and used to estimate the effect of changes. Other measures of central tendency might better reflect the typical time taken because the distributions are asymmetrical, but such measures cannot be used arithmetically to quantify the effect of changes on the required staff time. Because of the large sample, according to the central limit theorem estimates and inferences, including 95% confidence intervals, are likely to be robust because sample means are likely to be normally distributed.

## CONCLUSIONS

This is the first multi-site study to use a robust and validated methodology to investigate the nursing time involved in taking a set of vital signs observations and we provide the most reliable estimates of the time required. Our estimates show that the associated workload can be considerable. When observations are repeated across multiple patients on multiple occasions, the workload is even more significant. Therefore, any changes in observation frequency have potentially large implications for nursing resources that need to be considered, as well as the potential opportunity costs if observations are given higher priority than other nursing activities.

## Data Availability

Data are available upon request.

## ACKNOWLEDGMENTS

We would like to thank all the Local Principal Investigators at the Trusts for facilitating this study: Jody Ede, Karen Hill, Katrina Kennedy, Joanna Mouland.

## Footnotes

1* Level 0 care: patients whose needs can be met through normal ward care in an acute hospital and patients at risk of their condition deteriorating Level 1 care: patients recently relocated from higher levels of care, whose needs can be met on an acute ward with additional advice and support from the critical care team INTENSIVE CARE SOCIETY 2002. Levels of critical care for adult patients.

