## Supplemental material for "How long do nursing staff take to measure and record patients’ vital signs observations in hospital? A time-and-motion study"

Table S1	Mean length and 95% CI by staff type - raw estimates from rounds

| **Staff type** | **Including interruptions,  mean [95% CI]** | **Excluding non-vital-sign**  **interruptions,  mean [95% CI]** | **Excluding all interruptions,  mean [95% CI]** |
| --- | --- | --- | --- |
| All | 6:25 [6:04-6:47] | 5:00 [4:45-5:15] | 4:15 [4:06-4:26] |
| HCA | 6:10 [5:33-6:51] | 4:48 [4:24-5:14] | 4:11 [3:55-4:28] |
| RN | 6:32 [6:00-7:07] | 5:01 [4:40-5:24] | 4:14 [3:59-4:29] |
| STUDENT NURSE | 6:30 [5:49-7:15] | 5:12 [4:41-5:47] | 4:24 [4:02-4:48] |

Figure S1	Distribution of raw estimates from rounds

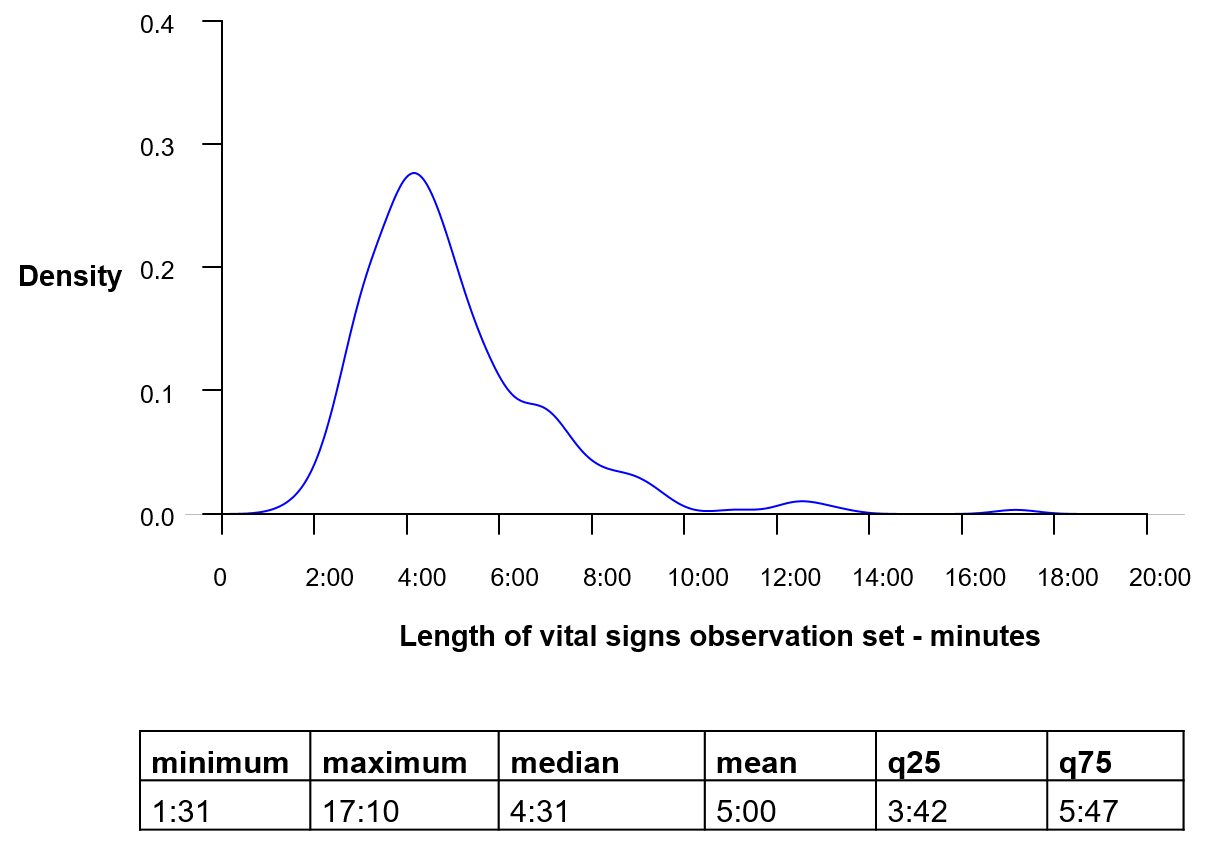

Mean (minutes:seconds): 5:00

Median (minutes:seconds): 4:31

Table S2	Mean length and 95% CI by staff type - raw estimates from time at the patient bedside only

| **Staff type** | **Including interruptions,  mean [95% CI]** | **Excluding non-vital-sign**  **interruptions,  mean [95% CI]** | **Excluding all interruptions,  mean [95% CI]** |
| --- | --- | --- | --- |
| All | 4:24 [4:15-4:33] | 3:45 [3:37-3:52] | 3:31 [3:25-3:37] |
| HCA | 4:21 [4:08-4:37] | 3:43 [3:32-3:55] | 3:33 [3:24-3:42] |
| RN | 4:22 [4:08-4:36] | 3:44 [3:32-3:57] | 3:29 [3:20-3:38] |
| STUDENT_NURSE | 4:34 [4:16-4:53] | 3:49 [3:35-4:05] | 3:34 [3:22-3:47] |

Figure S2	Distribution of raw estimates from time at the patient bedside only

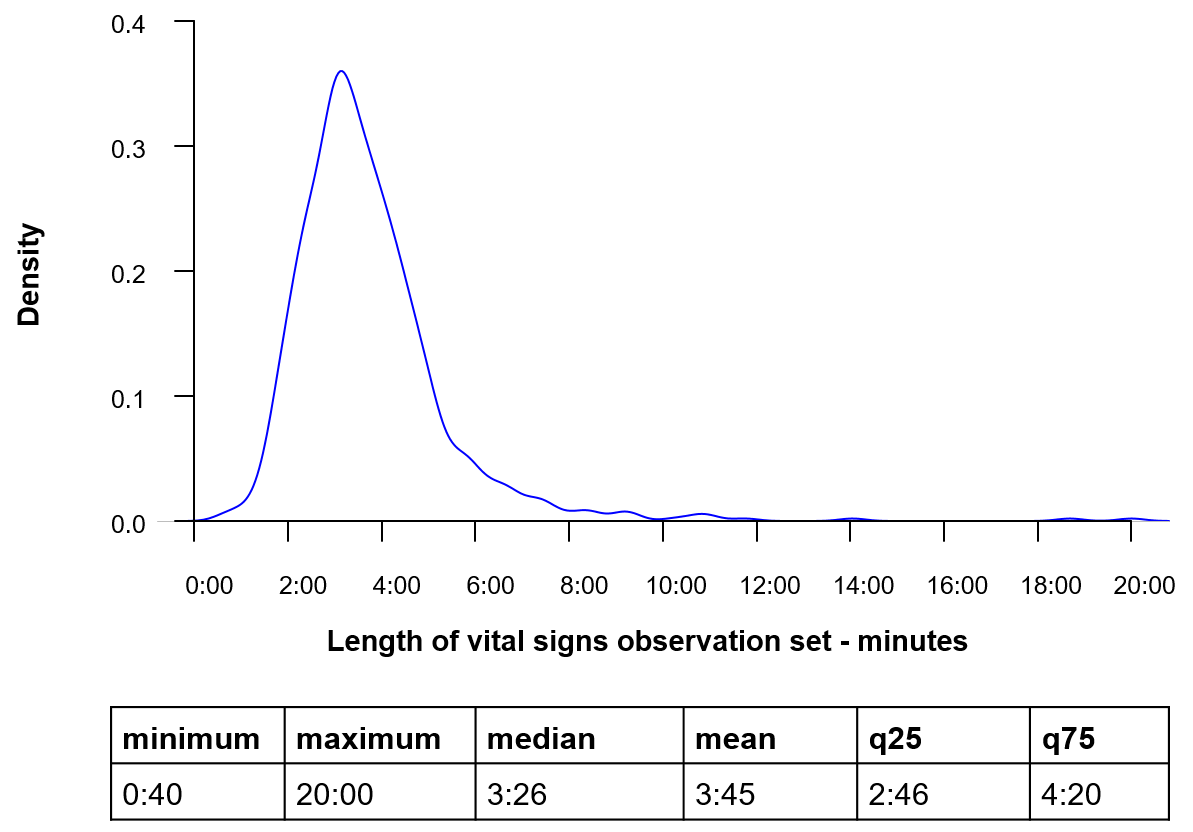

Mean (minutes:seconds): 3:45

Median (minutes:seconds): 3:26
